# Digital proximity tracing app notifications lead to faster quarantine in non-household contacts: results from the Zurich SARS-CoV-2 Cohort Study

**DOI:** 10.1101/2020.12.21.20248619

**Authors:** Tala Ballouz, Dominik Menges, Helene E Aschmann, Anja Domenghino, Jan S Fehr, Milo A Puhan, Viktor von Wyl

**Affiliations:** Epidemiology, Biostatistics and Prevention Institute, University of Zurich, Zurich, Switzerland; Department of Infectious Diseases and Hospital Epidemiology, University Hospital Zurich, University of Zurich, Zurich, Switzerland; Cantonal Health Directorate Zurich, Zurich, Switzerland; Department of Visceral and Transplantation Surgery, University Hospital Zurich, Zurich, University of Zurich, Switzerland; Institute for Implementation Science in Health Care, University of Zurich, Zurich, Switzerland

## Abstract

**Background:** Digital proximity tracing (DPT) apps may warn exposed individuals faster than manual contact tracing (MCT), leading to earlier interruption of transmission chains through quarantine. However, it is yet unclear whether these apps lead to a reduction in transmissions under real-world conditions. This study aimed at evaluating whether the SwissCovid DPT app is effective in warning close contacts of SARS-CoV-2 infected cases and prompting them to quarantine earlier.

**Methods:** A population-based sample of adult index cases and close contacts identified through MCT and enrolled in the Zurich SARS-CoV-2 Cohort study were surveyed regarding use of the SwissCovid app and SARS-CoV-2 exposure setting. We analyzed ooutcomes related to app effectiveness and adherence (i.e., receipt and uploading of notification codes by index cases; receipt of app warnings and steps taken by close contacts). Furthermore, we performed adjusted time-to-event analyses stratified by exposure setting to estimate the effect of the app on time between relevant exposure and entering quarantine among close contacts.

**Findings:** We included 393 index cases and 261 close contacts in the analysis. Among index cases using SwissCovid, 88% reported receiving and uploading a notification code in the app to trigger a warning among proximity contacts. Among close contacts using the app, 38% reported receiving an app warning due to the risk exposure. We found that non-household contacts who were notified by the app started quarantine at a median of 2 days after exposure, while those not notified started quarantine at a median of 3 days. In stratified multivariable analyses, app notified contacts had a greater probability of going into quarantine earlier than those without app notification (HR 1·53, 95% CI 1·15-2·03).

**Interpretation:** Our study showed that non-household contacts notified by the app started quarantine one day earlier than those not notified by the app. These findings constitute the first evidence that DPT may reach exposed contacts faster than MCT, leading to earlier quarantine and potential interruption of SARS-CoV-2 transmission chains.

**Funding:** Cantonal Health Directorate Zurich, University of Zurich Foundation and the Swiss Federal Office of Public Health.

## Background

Contact tracing is a crucial public health measure for controlling the spread of severe acute respiratory syndrome coronavirus 2 (SARS-CoV-2).^1,2^ Traditionally, contact tracing involves interviewing all infected individuals (index cases) to systematically identify their close contacts. This aims at interrupting viral transmission chains by referring these close contacts to quarantine and SARS-CoV-2 testing.^2–4^ However, such manual contact tracing (MCT) has inherent time delays, is resource intensive, and is limited by imperfect recall of encounters, especially those occurring briefly or by chance. Given the rapid SARS-CoV-2 transmission and the high proportion of asymptomatic cases, MCT alone is thus unlikely to be sufficiently effective.^5,6^ Digital proximity tracing (DPT) has been developed as a scalable complementary method to identify transmission chains that are likely to be missed or identified late by MCT.^7,8^

DPT applications register proximity encounters between individuals using the app with the aim to identify individuals that may have been exposed to an index case. Different technologies and architectures for DPT exist, one of which is decentralized, privacy-preserving proximity tracing (DP-3T) using Bluetooth Low Energy signals.^9^ The Swiss DPT app, «SwissCovid», was one of the first to be launched in June 2020 and follows the DP-3T blueprint. In November 2020, SwissCovid had around 1·84 million active users, corresponding to 21·9% of the Swiss population.^10^ Details on the design and implementation of SwissCovid are reported elsewhere.^8,11^

DPT has three potential advantages over MCT.^11^ First, the notification of exposed DPT app users is automatized once the index case has triggered the notification, leading to a potential speed advantage over MCT in interrupting transmission chains. Second, DPT still functions when MCT is at capacity due to high case numbers. Third, DPT has a wider reach than MCT because it does not rely on the infected individual’s recollection of his or her encounters. However, DPT apps are complex interventions involving multiple, sequential steps and specific actions by app users to exert their effect (notification cascade^12^, appendix 1). To date, the possible impact of DPT apps on pandemic mitigation is only partially understood. Some modeling studies reported that DPT, alone or in combination with MCT, can have an effect in reducing SARS-CoV-2 transmission.^7,13,14^ However, they relied on several strong assumptions, indicating that DPT effectiveness strongly depends on population uptake and the timeliness of case identification and quarantining of contacts.^15,16^ Furthermore, a recent analysis on the impact of the “Test-and-Trace” programme on the Isle of Wight, which includes a DPT app among other public health measures, demonstrated that it led to a marked decrease in SARS-CoV-2 incidence. However, the contribution of the DPT app in this programme was difficult to evaluate.^17^

Only few empirical analyses on the effectiveness of SwissCovid exist. Two studies identified factors associated with DPT app uptake and reasons for non-use^18^, as well as challenges related to the implementation of SwissCovid in Switzerland.^19^ Salathé et al. analyzed publicly available performance indicators for SwissCovid, such as number of app downloads and notification codes (CovidCodes) entered, and demonstrated proof-of-principle for the functioning of the app.^20^ Furthermore, a detailed quantification of the different steps of the SwissCovid notification cascade for the Canton of Zurich suggests that DPT may have led to an additional 5% of exposed persons entering quarantine in September 2020.^12^

Yet, critical questions pertaining to other conditions necessary for the functioning of DPT apps and their real-world impact remain unanswered. In particular, no data is available on the adherence of index cases and close contacts to recommended actions upon testing positive for SARS-CoV-2 (i.e., uploading of codes) or receiving an app notification (i.e., quarantine and testing). Furthermore, it is unclear whether the app indeed reduces the time between exposure and entering quarantine in close contacts. We aimed to fill this evidence gap in the Zurich SARS-CoV-2 Cohort study by addressing two main questions. First, we evaluated the adherence of SwissCovid app users with the recommended steps. Second, we examined the effectiveness of SwissCovid by evaluating whether the time from exposure to quarantine differed between close contacts who have or have not received an app warning.

## Methods

### Study design and participants

The Zurich SARS-CoV-2 Cohort Study is an ongoing, prospective, longitudinal, population-based cohort study of individuals infected with SARS-CoV-2 and their close contacts in the Canton of Zurich. The cohort was established in collaboration with the Cantonal Health Directorate Zurich and aims to characterize clinical outcomes and immunological responses of index cases and examine patterns of transmission among index cases and their close contacts.

Individuals diagnosed with SARS-CoV-2 infection and their close contacts were identified through mandatory laboratory reporting of positive cases to and routine contact tracing by the Cantonal Health Directorate. All identified index cases and close contacts were screened for eligibility and invited if they were ≥18 years old, residing in the Canton of Zurich, had sufficient knowledge of the German language and were able to follow the study procedures. Random sampling of the two populations was performed on a daily basis. Sampling of index cases was stratified by age and close contacts were sampled in clusters based on the respective index case. Informed consent was obtained from all individuals agreeing to participate in the study. In this analysis, we used data from index cases and close contacts enrolled between 07 August 2020 and 30 September 2020, when conditions changed due to a sharp increase in case numbers in Switzerland in early October 2020.^21^

The study protocol was approved by the ethics committee of the Canton of Zurich (BASEC 2020-01739) and prospectively registered on the International Standard Randomised Controlled Trial Number Registry (<u>ISRCTN14990068</u>).

### Data collection

Data was collected and managed through the Research Electronic Data Capture (REDCap) system. Questionnaires for index cases included questions on socio-demographics, comorbidities, details on the suspected transmission event, symptoms and disease burden. Similarly, questionnaires for close contacts elicited information regarding socio-demographics, symptoms, experiences with quarantine, and details on their contact with the index case (e.g., exposure setting, timing). Both questionnaires included questions related to the use of the SwissCovid app, receipt and uploading of CovidCodes by index cases, and app warnings received by close contacts (appendix 2).

Data from both questionnaires was available for ten individuals who were initially enrolled as close contacts and later tested SARS-CoV-2 positive. Individuals identified as close contacts by contact tracing who tested positive before study enrollment were directly enrolled as index cases and thus provided data from only one questionnaire (n=55; appendix 3). Individuals pertaining to either group were considered “converted index cases” in the analysis and contributed data on the level of close contacts and index cases, as appropriate.

### Definitions

Participants reporting permanent or occasional use of SwissCovid were considered app users. Self-reported exposure settings were classified as household if the participant reported living in the same household as the index case. Non-household settings included workplace, private settings, public settings, healthcare facility, school or university, shared accommodation, and military. In line with the definition by the Cantonal Health Directorate, the exposure date referred to the last day when the close contact was within 1.5 m distance of the index case for ≥15 minutes up to 48 hours before symptom onset (or positive test if asymptomatic) and without personal protective equipment. For household contacts, exposure date corresponded to the first day the index case was isolated. Exposure dates were recorded by two methods: self-reported by participants (main measure) and a proxy measure defined as 10 days prior to the last day of quarantine, as determined by contact tracing.

### Outcomes

To evaluate adherence, primary outcomes included the frequency of index cases who received and uploaded the CovidCode (thereby triggering a warning of contacts), frequency of close contacts who received a SwissCovid app notification and among those, and the frequency of close contacts who received the notification before being contacted by MCT. Regarding effectiveness, our primary outcome was the time interval (in days) between exposure date and the beginning of quarantine among close contacts, comparing those notified by the app to those not notified by the app.

### Statistical methods

Adherence with recommended actions was evaluated using descriptive statistics. Continuous variables are presented as median and interquartile ranges (IQR) and categorical variables as frequencies (N) and percentages (%). Free text responses regarding reasons for non-use of the app, not uploading the CovidCode by index cases and steps taken by close contacts after receiving a warning, were reviewed. Based on their context, responses were coded without a preconceived categorization and reported in frequency and percentage.

To evaluate effectiveness (i.e., time from exposure to beginning of quarantine), close contacts were grouped into “app notified” and “not app notified”. App non-users were considered “not app notified”. We assumed that notification time in household settings would be intrinsically faster than in non-household settings due to differences in information pathways and thus stratified participants by exposure setting (household vs. non-household). Concordance between self-reported and proxy exposure dates was examined. If self-reported date of last exposure was later than the beginning of quarantine (e.g., in same household contacts where the exposure date was not clearly defined), we used the proxy exposure date. If contacts entered quarantine on the day of exposure (leading to a 0 day interval) a delay of 0·5 days was added. Differences between groups were explored using Kaplan-Meier curves and stratified log-rank test. To evaluate the association between app notification and time from exposure to quarantine, we used a Cox proportional hazards model stratified by exposure setting and adjusted for age group, sex, education and employment status. Non-proportionality and possible influential outliers were tested using the scaled Schoenfeld residuals and *dfbeta* values, respectively. The model was adjusted for the cluster effect of sampling using robust variance estimation. Hazard ratios (HRs) and 95% confidence intervals (CI) were reported. We explored the robustness of our findings by performing a sensitivity analysis using the proxy exposure date instead of the self-reported exposure date to estimate the time from exposure to quarantine. Furthermore, in a second sensitivity analysis, we restricted our analysis to those using the app to account for potential confounding mediated by app use and associated characteristics of the close contacts. All analyses were performed using R version 3.6.1.

## Results

We included 328 index cases, 65 index cases that converted from originally being traced as a close contact and 261 close contacts (appendix 3). Index cases and close contacts were largely similar with respect to socio-demographic characteristics (Table 1). Median age of index cases and close contacts at time of identification was 38 and 35 years, respectively. Approximately 50% of the participants in both groups were female. Other characteristics such as Swiss nationality (79% and 84%), level of education (55% and 62% with a university or technical college degree), employment status (81% and 80% employed) and self-reported comorbidities (22% and 23% with at least one comorbidity) were also comparable between close contacts and index cases. Converted index cases were slightly different from the other two groups, with approximately 54% being female and 92% Swiss nationals.

**Table 1.** Baseline characteristics of the study population.

| Variable | Close contact, N = 261 | Converted index case, N = 65 | Index case, N = 328 |
| --- | --- | --- | --- |
| Age, years* | 35 (28, 51) | 39 (29, 55) | 38 (29, 51) |
| **Sex** |  |  |  |
| Female | 128/261 (49.0%) | 35/65 (53.8%) | 164/328 (50.0%) |
| Male | 133/261 (51.0%) | 30/65 (46.2%) | 164/328 (50.0%) |
| **Education** |  |  |  |
| Up to mandatory school | 9/257 (3.5%) | 4/65 (6.2%) | 13/327 (4.0%) |
| Vocational training/specialized<br>baccalaureate | 89/257 (34.6%) | 22/65 (33.8%) | 133/327 (40.7%) |
| Technical college or university studies | 159/257 (61.9%) | 39/65 (60%) | 181/327 (55.3%) |
| (Missing) | 4 | 0 | 1 |
| **Employment status** |  |  |  |
| Employed | 205/256 (80.1%) | 53/64 (82.8%) | 265/327 (81.0%) |
| Student | 33/256 (12.9%) | 6/64 (9.4%) | 27/327 (8.3%) |
| Unemployed | 18/256 (7.0%) | 5/64 (7.8%) | 35/327 (10.7%) |
| (Missing) | 5 | 1 | 1 |
| **Monthly household income (Swiss**<br>**Francs)** |  |  |  |
| <6000 | 87/247 (35.2%) | 21/62 (33.9%) | 111/314 (35.4%) |
| 6000-12000 | 101/247 (40.9%) | 32/62 (51.6%) | 120/314 (38.2%) |
| >12000 | 59/247 (24.9%) | 9/62 (14.5%) | 83/314 (26.4%) |
| (Missing) | 14 | 3 | 14 |
| **Nationality** |  |  |  |
| Swiss | 220/261 (84.3%) | 60/65 (92.3%) | 260/328 (79.3%) |
| Non-Swiss | 41/261 (15.7%) | 5/65 (7.7%) | 68/328 (20.7%) |
| **Chronic comorbid conditions** |  |  |  |
| At least one self-reported comorbid<br>chronic condition | 57/252 (23%) | 13/61 (21.3%) | 69/318 (21.7%) |
| No self-reported chronic comorbid<br>conditions | 195/252 (77%) | 48/61 (78.7%) | 249/318 (78.3%) |
| (Missing) | 9 | 4 | 10 |
| **Known exposure setting** |  |  |  |
| Knows or has strong suspicion | 253/257 (98.4%) | 60/65 (92.3%) | 152/328 (46.3%) |
| No | 4/257 (1.6%) | 5/65 (7.7%) | 176/328 (53.7%) |
| (Missing) | 4 | 0 | 0 |
| **Exposure setting (among those with known/suspected exposure)** |  |  |  |
| Household | 72/252 (28.6%) | 32/60 (53.4%) | 22/150 (14.6%) |
| Workplace | 43/252 (17.1%) | 2/60 (3.3%) | 23/150 (15.3%) |
| Private setting | 66/252 (26.2%) | 15/60 (25.0%) | 42/150 (28.0%) |
| Public space | 45/252 (17.9%) | 8/60 (13.3%) | 43/150 (28.7%) |
| School/University | 8/252 (3.2%) | 0/60 (0.0%) | 1/150 (0.7%) |
| Other | 18/252 (7.0%) | 3/60 (5.0%) | 19/150 (12.7%) |
| Healthcare facility | 0/252 (0.0%) | 0/60 (0.0%) | 0/150 (0.0%) |
| (Missing) | 1 | 0 | 2 |
| **SwissCovid app use** |  |  |  |
| App non-user | 73/258 (28.3%) | 22/64 (34.4%) | 125/326 (38.3%) |
| App user | 185/258 (71.7%) | 42/64 (65.6%) | 201/326 (61.7%) |
| (Missing) | 3 | 1 | 2 |
| **Reasons for non-use of the app** |  |  |  |
| No knowledge of the app | 1/57 (1.8%) | 2/16 (12.5%) | 5/104 (4.8%) |
| Perception of uselessness | 13/57 (22.8%) | 3/16 (18.8%) | 29/104 (27.9%) |
| Technical problems | 14/57 (24.5%) | 4/16 (25.0%) | 21/104 (20.2%) |
| Privacy and data protection | 16/57 (28.1%) | 5/16 (31.2%) | 23/104 (22.1%) |
| Other | 13/57 (22.8%) | 2/16 (12.5%) | 26/104 (25.0%) |
| (Missing) | 16 | 6 | 22 |

\*Median (IQR)

The exposure setting was reported as known or strongly suspected by 98% of close contacts and 93% of converted index cases. Meanwhile, only 46% of index cases knew or suspected the setting in which SARS-CoV-2 transmission occurred. Among those with knowledge or suspicion regarding their exposure, household and private settings were most frequently reported among close contacts (29% and 26%) and converted index cases (53% and 26%). Index cases most frequently stated public spaces (29%) and private settings (28%) as the exposure setting.

### Adherence

Overall, 62% (n=201) of index cases, 66% (n=42) of converted participants, and 72% (n=185) of close contacts were app users. Reasons for app non-use are reported in Table 1 and appendix 4. On average, app non-users were older and a higher proportion were female, retired and non-Swiss nationals compared to app users (appendix 5).

Among 243 index cases using SwissCovid, 92% (n=224) reported to have received a CovidCode from public health authorities. Of those, 96% (n=215) uploaded the code in the app, thus triggering a notification to potentially exposed contacts. Main reasons for not uploading the code included receiving it too late or that their close contacts were already in quarantine (Table 2).

**Table 2:**
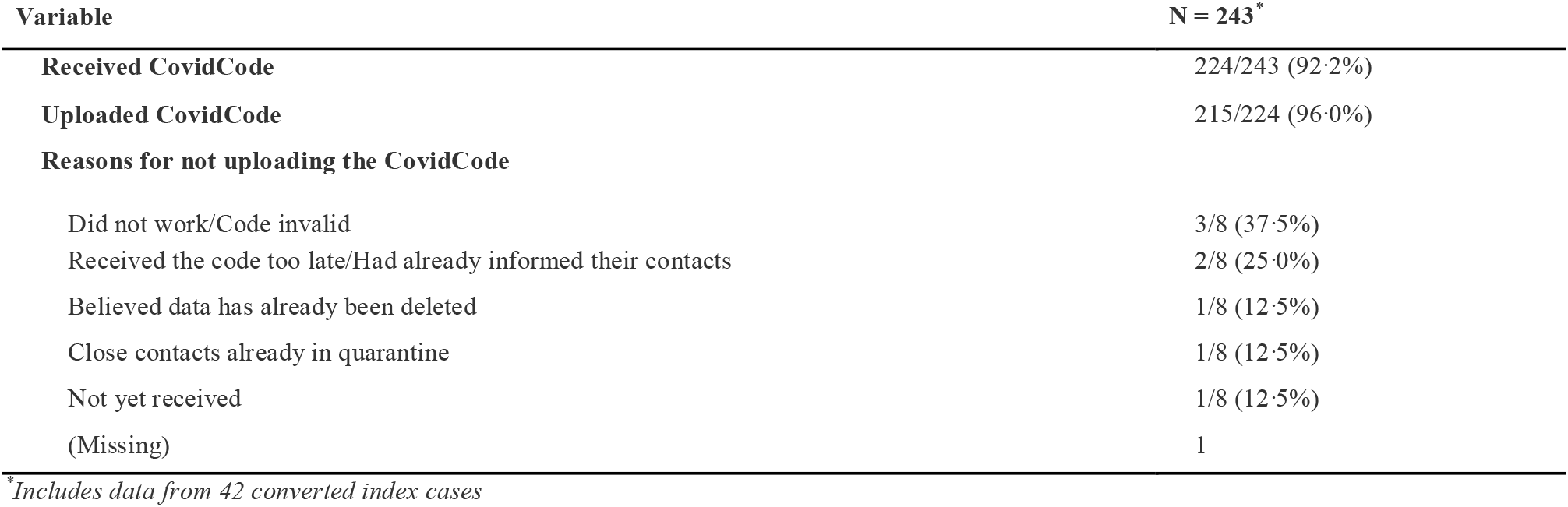
CovidCodes received and uploaded by index cases who are app users.

| Variable | N = 243* |
| --- | --- |
| **Received CovidCode** | 224/243 (92.2%) |
| **Uploaded CovidCode** | 215/224 (96.0%) |
| **Reasons for not uploading the CovidCode** |  |
| Did not work/Code invalid | 3/8 (37.5%) |
| Received the code too late/Had already informed their contacts | 2/8 (25.0%) |
| Believed data has already been deleted | 1/8 (12.5%) |
| Close contacts already in quarantine | 1/8 (12.5%) |
| Not yet received | 1/8 (12.5%) |
| (Missing) | 1 |

\*Includes data from 42 converted index cases

Among the 192 close contacts using the app, 38% (n=73) received an app notification within 7 days of the last relevant exposure. Out of these, 12% (n=9) received the notification before being contacted by MCT. After receiving the app notification, 14% of the 73 close contacts followed the recommendation of calling the SwissCovid info-line, whilst the remainder undertook other (19%) or no actions (67%). Most participants taking no action stated that they had already been reached by MCT and were already in quarantine and/or tested for SARS-CoV-2 (Table 3).

**Table 3:** App notifications received and steps taken after by close contacts who are app users.

| Variable | Close contacts; N = 192* |
| --- | --- |
| **Received a notification by the app** |  |
| Yes, in the last 7 days probably because of the current contact | 73/192 (38.0%) |
| Yes, more than 7 days ago | 2/192 (1.0%) |
| No notification | 117/192 (60.9%) |
| **Notified by the app before the cantonal medical service** | 9/73 (12.3%) |
| (Missing) | 2 |
| **Steps taken after receiving an app notification** |  |
| Called SwissCovid infoline | 10/72 (13.9%) |
| Other steps taken | 14/72 (19.4%) |
| No steps taken | 48/72 (66.7%) |
| (Missing) | 3 |
| **Other steps taken** |  |
| Had already taken measure** after being traced by contact tracing | 6/14 (42.9%) |
| Had already taken measures** following family/friend's advice | 6/14 (42.9%) |
| Called cantonal medical service | 1/14 (7.1%) |
| Testing | 1/14 (7.1%) |

\*Includes 7 "Converted" cases for whom data as a close contact was available (i.e. converted after enrolment)
\*\*Includes SARS-CoV-2 testing and quarantine

### Effectiveness

The median time from last exposure to beginning of quarantine among all close contacts was 2 days (IQR 1-3 days) based on the self-reported exposure date (main analysis). When using the proxy exposure date, the median time to quarantine was 1 day (IQR 0·5-3 days, sensitivity analysis). There was a 69% concordance between self-reported and proxy exposure date. 20 close contacts reported to have had the last exposure after starting quarantine, 18 of which reported the index case to be a household member and two a friend.

We found that the time from exposure to quarantine differed across exposure settings and between contacts that received or did not receive an app notification (Figure 1). Overall, household contacts had a shorter median time from exposure to quarantine than non-household contacts (1 day vs. 3 days). In *non-household* settings, we found a difference in time intervals indicating a shorter duration to quarantine in app notified (n=43; median 2 days, IQR 1-3) compared to non-app notified contacts (n=138; median 3 days, IQR 2-4; p=0·01). Among the 43 app notified non-household contacts, 8 (18·6%) reported to have received the app notification before they were contacted by MCT. 47% of app notified non-household contacts reported to have decided themselves to initiate quarantine compared to 31% of non-app notified non-household contacts. In app notified contacts that received the warning before MCT, 75% (6/8) reported self-quarantine as the initial reason for quarantine, compared to 41% (14/34) of those receiving the warning after MCT (appendix 6). However, in *household* settings, there was no evidence for a difference in the time from exposure to quarantine between app notified (median 0·5 days, IQR 0·5-2·0) and non-app notified contacts (median 1 day, IQR 0·5-2·0; p=0·11).

**Figure 1:**
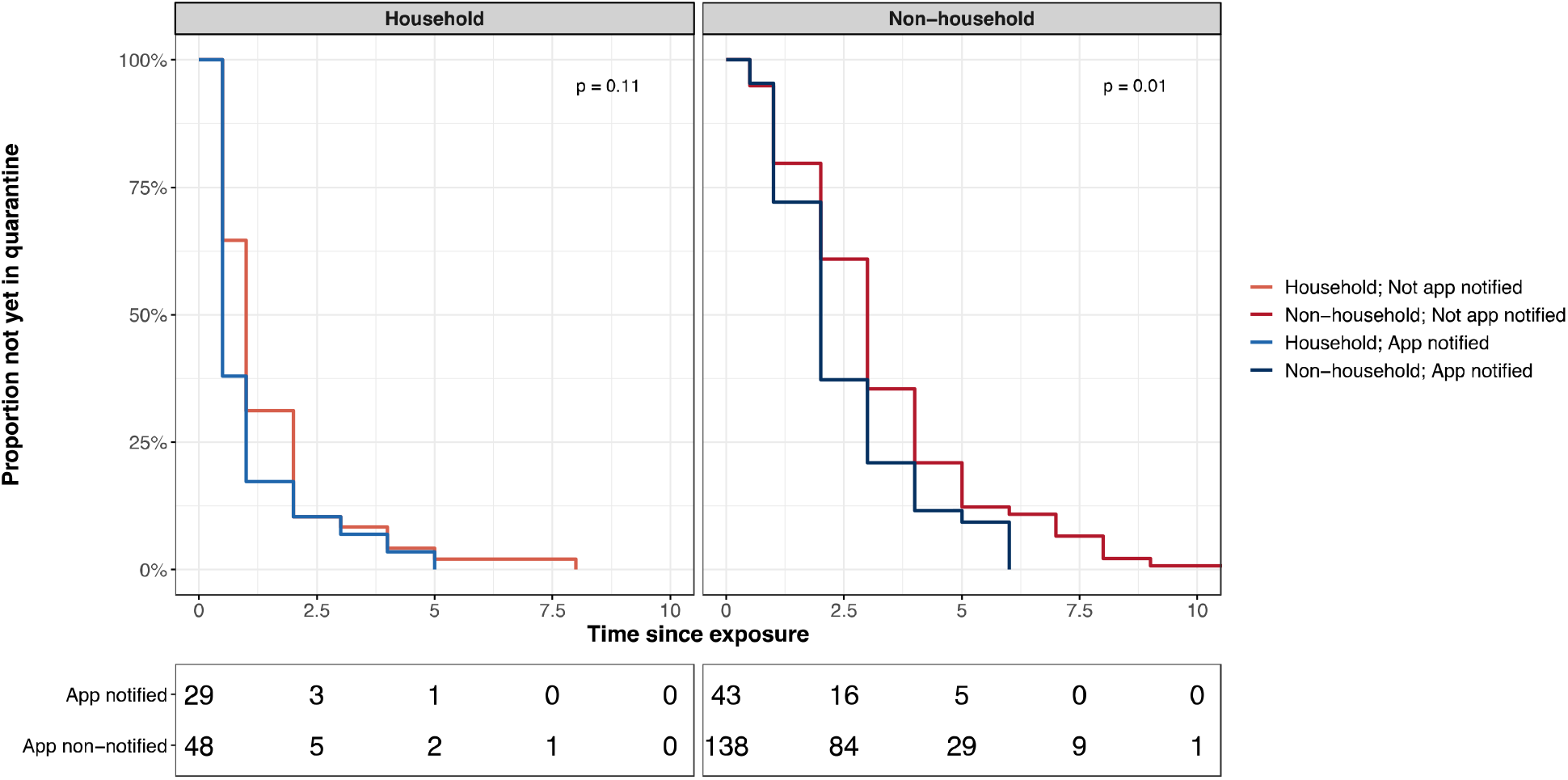
Time from exposure to quarantine in app notified versus not app notified stratified by exposure setting.

In the stratified multivariable Cox model, we found strong evidence that contacts notified by the app had a greater probability of going into quarantine earlier than those not notified by the app while adjusting for age, sex, education, and employment status (HR 1·53, 95% CI 1·15-2·03; p=0·004). Age, education level, and employment status were not associated with a shorter time to quarantine (Table 4, appendix 7). No interaction was detected between app notification and exposure setting. Sensitivity analyses using the proxy exposure date, as well as when restricting the analysis to app users, yielded similar results (appendices 8 and 9).

**Table 4.** Multivariable cox proportional hazards analysis of time from exposure to quarantine in close contacts, stratified by exposure setting.

| Variable | AHR (95% CI) * | p-value |
| --- | --- | --- |
| **Age at diagnosis, years** |  |  |
| 18-39 | 1 (Reference) |  |
| 40-64 | 1.28 (0.91 – 1.80) | 0.16 |
| 65+ | 1.45 (0.67 – 3.12) | 0.35 |
| **Sex** |  |  |
| Female | 1 (Reference) |  |
| Male | 0.86 (0.63 – 1.15) | 0.31 |
| **Highest education level** |  |  |
| Mandatory school | 1 (Reference) |  |
| Vocational training/ baccalaureate | 0.67 (0.34 – 1.32) | 0.24 |
| Technical college or university | 0.61 (0.29 – 1.27) | 0.19 |
| **Employment status** |  |  |
| Employed | 1 (Reference) |  |
| Student | 1.41 (0.91 – 2.18) | 0.12 |
| Unemployed | 0.85 (0.38 – 1.93) | 0.70 |
| **App notification** |  |  |
| App non-notified | 1 (Reference) |  |
| App notified | 1.53 (1.15 – 2.03) | 0.004 |

AHR: adjusted hazard ratio, CI: confidence interval \*Model adjusted for age group, sex, highest education level, employment status and app notification

## Discussion

In this study of 261 close contacts and 393 index cases (including 65 converted individuals) identified through routine contact tracing in the Canton of Zurich, we evaluated use of SwissCovid and whether it provides a time advantage over MCT. Our analysis showed that non-household contacts notified by the app started quarantine earlier than those not notified by the app. This provides important evidence that DPT apps have an impact on the timely interruption of transmissions chains.

Most household contacts entered quarantine the same or the following day after exposure to an index case in our study. This was expected, as they are easier to contact and are commonly informed directly by the index case about their exposure. On the other hand, the contacting of non-household contacts through MCT is often more time-consuming and longer delays may be expected. We found evidence for a possible time advantage through the app in non-household setting, with app-notified contacts entering quarantine on average one day earlier than those not notified by the app. Considering that the testing delay (i.e., time from symptom onset to positive test) is 2.5 days on average in the Canton of Zurich^22^, tracing delays and an overall reduced effectiveness of a contact tracing strategy are to be expected.^13^ However, this does not explain differences between app notified and not app notified contacts. To explain this difference, we descriptively explored multiple hypotheses. We found that a higher percentage of app notified non-household contacts reported to have entered self-quarantine compared to those not notified by the app (47% vs. 31%). This finding supports the hypothesis that receiving an app notification may lead to a shorter time between exposure and quarantine. Although only 8 (19%) of 43 app notified contacts received the app warning before being reached by MCT, app notifications received after being called by contact tracers may not be without effect. For example, these notifications could have a reinforcing effect on the quarantine recommendations by MCT. We additionally explored alternative hypotheses that could explain our findings, such as confounding by index case characteristics (e.g., more symptomatic cases or earlier testing among app users) or differences in health literacy. We found no indication for systematic confounding in our descriptive analyses. However, more in-depth studies are clearly warranted.

Our findings constitute the first evidence that DPT may be effective in reaching close contacts faster than MCT. Albeit small, such a time difference may be relevant in reducing transmission in the population. Ferretti et al. demonstrated in a modeling study that reducing the time to quarantine from 3 to 2 days had a substantial impact on reducing the spread of SARS-CoV-2, assuming that a large fraction of the population is using the app.^7^ This emphasizes the need to focus on behavioral aspects of app uptake and use for the implementation of DPT, as well as to consider specific subgroups in its evaluation, such as distinguishing between different exposure settings.

In our study, participants were enrolled during a period when case numbers were comparatively low. One potential advantage of the app is that it may be even more effective in times when case numbers are high leading to capacity issues in MCT. However, this requires that app coverage is sufficient and an efficient process is in place to initiate the notification cascade.^12,20^ Thus, our findings may underestimate the effectiveness of the SwissCovid app in situations where MCT is overwhelmed. In addition, this analysis is restricted to close contacts that were identified by MCT, due to the design of the Zurich SARS-CoV-2 Cohort study. However, another potential advantage of the app is in warning exposed individuals that were unknown to the index case or about whom they had forgotten.^7^ This setting was impossible to evaluate due to the privacy-by-design principle implemented in the SwissCovid app. As a consequence, our analysis did not consider potential additional benefits arising in the context of such exposure events.

A high percentage of participants reported using the app, exceeding previous estimates based on publicly available data and other population-based surveys.^18,20^ This difference may be explained by participants enrolled in the Zurich SARS-CoV-2 Cohort study being better informed about COVID-19 and more compliant with preventive public health measures than non-participants. Based on our data, the generation and uploading of CovidCodes seemed efficient, which contrasts previous reports of an approximate 30% gap between generated and entered CovidCodes.^20^ Nevertheless, some of the participants also reported significant delays in receiving the CovidCodes, as also indicated by the relatively low proportion of close contacts being notified by the app before being reached by MCT. Until recently, CovidCodes have been issued only by cantonal medical services and provided to index cases at the initial contact by contact tracing team. However, additional steps have been taken to improve the efficiency of the processes necessary for reducing such delays. From 18 November 2020, health service providers such as laboratories, pharmacies and testing centers were able to generate and issue CovidCodes to index cases to ensure the more rapid initiation of the notification cascade resulting in the warning of exposed contacts. Furthermore, since 12 December 2020, CovidCodes are generated and sent automatically through an online form completed by index cases as the first step in MCT in the Canton of Zurich.

Some further limitations of our study should be noted. Selection effects during enrollment may have led to a generally more health literate or compliant study population. However, such self-selection effects would not invalidate the proof-of-principle of our analysis but limit the transportability of our findings to the general population. Furthermore, despite consistent signals in our data, a causality between app notification and faster quarantine could not be unequivocally demonstrated. But the observation of a small subgroup of contacts who received the app notification and entered quarantine before being reached by MCT instills confidence that SwissCovid, in principle, achieves one of its main goals. Further studies are needed to quantify the impact of our findings on pandemic mitigation.

To our knowledge, our study is the first to evaluate the real-world effectiveness of a DPT app and leverages data from a prospective population-based cohort study. While a more in-depth assessment of the exact sequence and timing of events related to the notification cascade may shed further light on the impact of SwissCovid on an individual level, our findings confirm the hypothesized benefit of DPT apps alarming non-household contacts earlier than MCT, thereby leading to earlier quarantine.

## Data Availability

We are open to sharing individual participant data that underlie the results reported in this article, after de-identification upon reasonable requests to the corresponding author. Data requestors will need to sign a data access agreement.

## Contributors

DM, TB, HEA, JSF and MAP conceived and planned the Zurich SARS-CoV-2 Cohort study. DM, TB and MAP coordinated the the Zurich SARS-CoV-2 Cohort study. TB, DM, and VvW designed this study. HEA implemented and coordinated the sampling and recruitment of study participants. TB, DM, and AD contributed to the enrolment of study participants and data collection. TB and DM prepared the data and performed the statistical analysis. TB, DM and VvW drafted the first manuscript. All authors critically revised the draft manuscript and read and approved the final manuscript. MAP, JSF, and VvW acquired funding for the project.

## Acknowledgments

The Zurich SARS-CoV-2 Cohort study is part of the Corona Immunitas research program, coordinated by the Swiss School of Public Health (SSPH+) and funded through SSPH+ fundraising, including funding by the Swiss Federal Office of Public Health, the Cantons of Switzerland (Basel, Vaud and Zurich), private funders (ethical guidelines for funding stated by SSPH+ were respected) and institutional funds of the participating universities. Additional funding specific to this study was provided by the Cantonal Health Directorate Zurich, the University of Zurich Foundation and the Swiss Federal Office of Public Health. We would like to thank the Zurich SARS-CoV-2 administrative team for their great efforts and dedication to the study. We also thank all participants of the Zurich SARS-CoV-2 Cohort for their invaluable contribution to our study.

