## Supplementary Material for "Digital proximity tracing app notifications lead to faster quarantine in non-household contacts: results from the Zurich SARS-CoV-2 Cohort Study"

**Appendix 1**: **Steps of the notification cascade of the SwissCovid app**

The notification cascade of SwissCovid involves specific actions taken by index cases and close contacts. Whenever an app user tests positive for SARS-CoV-2 (index case), this person receives an activation code (CovidCode) from the public health authorities, which has to be uploaded in the app to trigger notifications to other app users. The code upload then leads to a warning of proximity contacts who were within close distance (<1.5 m) for 15 minutes or more during the time window of infectivity of the SARS-CoV-2 positive case. Upon app notification, exposed contacts are advised to call the SwissCovid infoline, get tested and enter quarantine. Although all these steps are voluntary, they are strongly encouraged.

**Appendix 2: SwissCovid app related questions in participant questionnaires**

I. **Index cases**

| 1. | Do you use the SwissCovid App? | - Yes, always - Yes, but sometimes I switch off Bluetooth to interrupt the SwissCovid app - No, I have de-installed it - No but I plan on using it - No |
| --- | --- | --- |
| 2. | If answer to 1 is any of the «No» options  Why are you currently not using the SwissCovid App? | - I do not know the app - I don't think that the app is useful for me - I cannot install the app (e.g. due to technical problems, because I don't have an Android or iOS smartphone) - I fear for my privacy and data protection - Other reasons (please specify) |
| 3. | If answer to 1 is any of the «Yes» options  Have you received a CovidCode (activation code that you receive from the cantonal authorities after a positive coronavirus test in order to warn other people via the app)? | - Yes - No |
| 4. | If 3= «Yes»  Have you entered the CovidCode in the SwissCovid App to activate the anonymous notification of other App users? | - Yes - No |
| 5. | If 4= «No»  Can you give the reason why you have not or could not activate the CovidCode? | Free text |

II. **Close contacts**

| 1. | Do you use the SwissCovid App? | - Yes, always - Yes, but sometimes I switch off Bluetooth to interrupt the SwissCovid app - No, I have de-installed it - No but I plan on using it - No |
| --- | --- | --- |
| 2. | If answer to 1 is any of the «No» options  Why are you currently not using the SwissCovid App? | - I do not know the app - I don't think that the app is useful for me - I cannot install the app (e.g. due to technical problems, because I don't have an Android or iOS smartphone) - I fear for my privacy and data protection - Other reasons (please specify) |
| 3. | If answer to 1 is any of the «Yes» options  Has the SwissCovid App ever issued a warning that you have been in contact with a person infected with the coronavirus? | - Yes, probably because of the current contact (i.e. in the last 7 days) - Yes, at an earlier date (i.e. more than 7 days ago) - Yes, probably because of the current contact as well as at an earlier time - No, I have not had any warning |
| 4. | If answer to 3 is 1st or 3rd option  Did you receive a warning from the SwissCovid App before you were contacted by the cantonal medical service? | - Yes - No |
| 5. | If answer to 4 is “Yes”  What steps did you take after you were warned by the app? | - I have called the recommended SwissCovid Infoline - I have taken other steps, as follows (please specify) - I have taken no further steps |

**Appendix 3: Study enrollment and populations in the Zurich SARS-CoV-2 Cohort study**

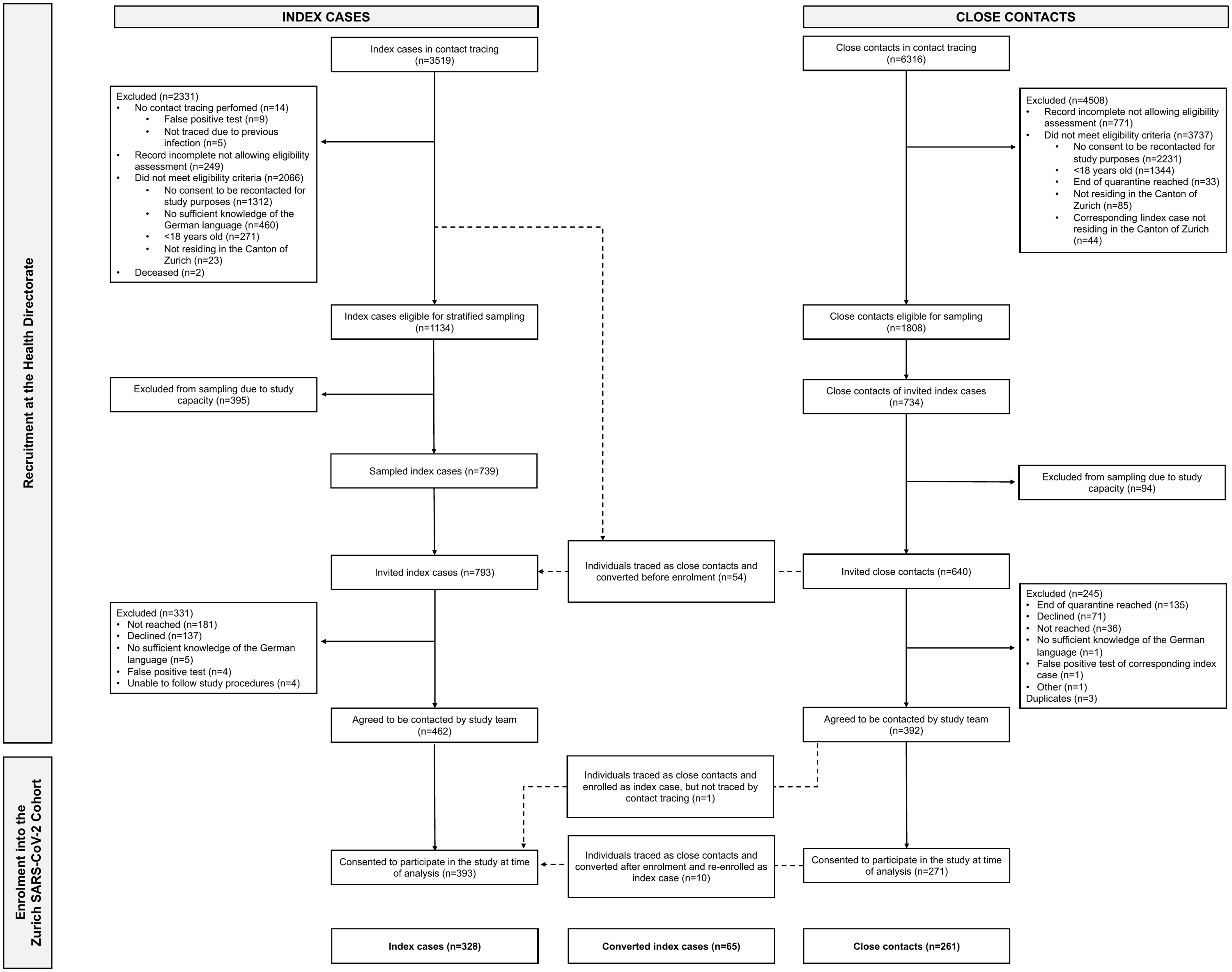

**Appendix 4: Other reasons for non-use of the SwissCovid app, as reported by participants**

| **Variable** | **Close contact, N = 13** | **Converted index case, N = 2** | **Index case, N = 26** |
| --- | --- | --- | --- |
| Consumes too much battery | 2/13 (15%) | 1/2 (50%) | 5/25 (20%) |
| Does not carry a cell phone often | 1/13 (8%) | 0/2 (0%) | 3/25 (12%) |
| Almost always has bluetooth off | 2/13 (15%) | 0/2 (0%) | 0/25 (0%) |
| Does not always have internet | 1/13 (8%) | 0/2 (0%) | 0/25 (0%) |
| App not working correctly | 1/13 (8%) | 0/2 (0%) | 0/25 (0%) |
| Data costs | 1/13 (8%) | 0/2 (0%) | 0/25 (0%) |
| Personal decision/No specific reason | 1/13 (8%) | 0/2 (0%) | 4/25 (16%) |
| Does not believe infected persons report | 0/13 (0%) | 0/2 (0%) | 0/25 (0%) |
| No confidence in the app | 1/13 (8%) | 0/2 (0%) | 0/25 (0%) |
| Privacy and does not have the right phone | 1/13 (8%) | 0/2 (0%) | 0/25 (0%) |
| Privacy and worries about false reports | 1/13 (8%) | 0/2 (0%) | 0/25 (0%) |
| Worried about the consequences of getting a notification (quarantine) | 0/13 (0%) | 0/2 (0%) | 2/25 (8%) |
| Feels constant stress/panic with the app | 0/13 (0%) | 1/2 (50%) | 2/24 (8%) |
| No longer recommended by the health department | 0/13 (0%) | 0/2 (0%) | 1/25 (4%) |
| Has to turn it off at work | 0/13 (0%) | 0/2 (0%) | 2/25 (8%) |
| Uses German corona app (border crosser) | 0/13 (0%) | 0/2 (0%) | 1/25 (4%) |
| (Missing) | 0 | 0 | 1 |

**Appendix 5: Baseline socio-demographic characteristics of app users and non-users**

|  | **Close contacts** | | **Converted index case** | | **Index case** | | **Overall** | |
| --- | --- | --- | --- | --- | --- | --- | --- | --- |
| **Variable** | **App non-user** N = 73 | **App user** N = 185 | **App non-user** N = 22 | **App user** N = 42 | **App non-user** N = 126 | **App user** N = 201 | **App non-user** N = 221 | **App user** N = 428 |
| **Age, years^*^** | 35 (26 – 55) | 35 (29 – 49) | 50 (31 – 62) | 38 (28 – 47) | 42 (29 – 54) | 37 (29 – 49) | 41 (28 – 55) | 36 (29 – 49) |
| **Sex** |  |  |  |  |  |  |  |  |
| Female | 39/73 (53·4%) | 88/185 (47·6%) | 15/22 (68.2%) | 19/42 (45·2%) | 69/126 (54·8%) | 94/201 (46·8%) | 123/221 (55·8%) | 201/428 (47·1%) |
| Male | 34/73 (46·6%) | 97/185 (52·4%) | 7/22 (31·8%) | 23/42 (54·8%) | 57/126 (45·2%) | 107/201 (53·2%) | 98/221 (44·2%) | 227/428 (52·9%) |
| **Education** |  |  |  |  |  |  |  |  |
| Mandatory school | 3/72 (4·2%) | 6/184 (3·3%) | 3/22 (13·6%) | 1/42 (2·4%) | 9/126 (7·1%) | 3/200 (1·5%) | 15/223 (6·7%) | 10/433 (2·3%) |
| Vocational training/baccalaureate | 41/72 (56·9%) | 48/184 (26·1%) | 9/22 (40·9%) | 13/42 (31·0%) | 53/126 (42·1%) | 80/200 (40·0%) | 103/223 (46·6%) | 141/433 (33·0%) |
| Technical college or university studies | 28/72 (38·9%) | 130/184 (70·7%) | 10/22 (45·5%) | 28/42 (66·6%) | 64/126 (50·8%) | 117/200 (58·5%) | 102/223 (46·6%) | 275/433 (64·7%) |
| (Missing) | 1 | 1 |  |  | 0 | 1 | 1 | 2 |
| **Employment status** |  |  |  |  |  |  |  |  |
| Employed | 54/72 (75·0%) | 150/183 (82·0%) | 18/22 (81·8%) | 35/41 (85·4%) | 96/126 (76·2%) | 168/200 (84·0%) | 168/223 (76·7%) | 353/431 (83·3%) |
| Student | 11/72 (15·3%) | 22/183 (12·0%) | 1/22 (4·5%) | 5/41 (12·2%) | 13/126 (10·3%) | 14/200 (7·0%) | 25/223 (11·2%) | 41/431 (9·7%) |
| Unemployed | 7/72 (9·7%) | 11/183 (6·0%) | 3/22 (13·6%) | 1/41 (2·4%) | 17/126 (13·5%) | 18/200 (9·0%) | 27/223 (12·1%) | 30/431 (7·0%) |
| (Missing) | 1 | 2 | 0 | 1 | 0 | 1 | 1 | 4 |
| **Nationality** |  |  |  |  |  |  |  |  |
| Swiss | 58/73 (79·5%) | 161/185 (87·0%) | 21/22 (95·5%) | 38/42 (90·5%) | 92/126 (73·0%) | 168/201 (83·6%) | 171/224 (77·7%) | 367/435 (86·0%) |
| Non-Swiss | 15/73 (20·5%) | 24/185 (13·0%) | 1/22 (4·5%) | 4/42 (9·5%) | 34/126 (27·0%) | 33/201 (16·4%) | 50/224 (22·3%) | 61/435 (14·0%) |
| **Chronic comorbidity** |  |  |  |  |  |  |  |  |
| At least one self-reported comorbid condition | 11/69 (15·9%) | 46/182 (25·3%) | 5/22 (22·7%) | 8/38 (21·1%) | 24/123 (19·5%) | 45/194 (23·2%) | 40/217 (18·9%) | 100/421 (23·8%) |
| No self-reported comorbid conditions | 58/69 (84·1%) | 136/182 (74·7%) | 17/22 (77·3%) | 30/38 (78·9%) | 99/123 (80·5%) | 149/194 (76·8%) | 174/217 (81·1%) | 321/421 (76·2%) |
| (Missing) | 4 | 3 | 0 | 4 | 3 | 7 | 7 | 14 |
| **Known exposure** |  |  |  |  |  |  |  |  |
| Knows or has strong suspicion | 72/73 (98·6%) | 181/184 (98·4%) | 21/22 (95·5%) | 38/42 (90·5%) | 66/126 (52·4%) | 85/201 (42·3%) | 159/224 (72·3%) | 311/434 (71·7%) |
| No | 1/73 (1·4%) | 3/184 (1·6%) | 1/22 (4·5%) | 8/42 (9·5%) | 60/126 (47·6%) | 116/201 (57·7%) | 62/224 (27·7%) | 123/434 (28·3%) |
| (Missing) | 0 | 1 |  |  |  |  | 0 | 1 |
| **Exposure setting** |  |  |  |  |  |  |  |  |
| Household | 19/71 (26·8%) | 53/181 (29·3%) | 12/21 (57·1%) | 20/38 (52·6%) | 13/66 (19·7%) | 8/83 (9·6%) | 44/161 (28·0%) | 85/309 (27·5%) |
| Non-household | 52/71 (73·2%) | 128/181 (70·7%) | 9/21 (42·9%) | 18/38 (47·4%) | 53/66 (80·3%) | 75/83 (90·4%) | 114/161 (72·0%) | 224/309 (72·5%) |
| (Missing) | 2 | 0 | 0 | 4 | 0 | 2 | 1 | 2 |

*^*^Median (IQR)*

| **Variable** | **Not app notified** | **App notified** | |
| --- | --- | --- | --- |
|  | **N = 141** | **Warned before MCT** **, N = 8** | **Warned after MCT, N = 34** |
| **Initial reason for initiating quarantine** |  |  |  |
| Contacted by GD | 67/141 (47·5%) | 2/8 (25·0%) | 15/34 (44·2%) |
| Self-quarantine^*^ | 43/141 (30·5%) | 6/8 (75·0%) | 14/34 (41·1%) |
| Employer/occupational health service instructions | 17/141 (12·1%) | 0/8 (0·0%) | 2/34 (5·9%) |
| Health care professional instructions | 1/141 (0·7%) | 0/8 (0·0%) | 0/34 (0·0%) |
| Followed family/friends advice | 4/141 (2·8%) | 0/8 (0·0%) | 2/34 (5·9%) |
| Other | 9/141 (6·4%) | 0/8 (0·0%) | 1/34 (2·9%) |
| *MCT: manual contact tracing*  *^*^Includes a SwissCovid app notification* MCT: manual contact tracing | | | |

**Appendix 6: Reasons for initiating quarantine in close contacts with non-household exposure, stratified by app notification status and timing of the warning (before or after manual contact tracing)**

**Appendix 7: Model including only those reporting non-household exposure setting**

| **Variable** | **AHR**  **(95% CI) ^*^** | **p-value** |
| --- | --- | --- |
| **Age at diagnosis, years** | |  |
| 18-39 | 1 (Reference) |  |
| 40-64 | 1·38 (0·90 – 2·12) | 0·14 |
| 65+ | 0·49 (0·14 – 1·71) | 0·007 |
| **Sex** |  |  |
| Female | 1 (Reference) |  |
| Male | 0·85 (0·58 – 1·24 | 0·39 |
| **Highest education level** | |  |
| Mandatory school | 1 (Reference) |  |
| Vocational training/ baccalaureate | 1·05 (0·56 – 1·97) | 0·87 |
| Technical college or university | 0·98 (0·46 – 1·79) | 0·78 |
| **Employment status** | |  |
| Employed | 1 (Reference) |  |
| Student | 1·92 (1·11 – 3·33) | 0·02 |
| Unemployed | 2·08 (0·79 – 5·47) | 0·14 |
| **App notification** | |  |
| Not app notified | 1 (Reference) |  |
| App notified | 1·53 (1·06 – 2·22) | 0·02 |
| *AHR: adjusted hazard ratio, CI: confidence interval*  *^*^Model adjusted for age group, sex, highest education level, employment status and app notification* | | |

**Appendix 8: Sensitivity analysis excluding close contacts not using the app**

***Model stratified by exposure setting***

| **Variable** | **AHR (95% CI) ^*^** | **p-value** |
| --- | --- | --- |
| **Age at diagnosis, years** | |  |
| 18-39 | 1 (Reference) |  |
| 40-64 | 1·38 (0·81 – 1·74) | 0·37 |
| 65+ | 2·5 (1·29 – 4·87) | 0·007 |
| **Sex** |  |  |
| Female | 1 (Reference) |  |
| Male | 0·77 (0·56 – 1·06) | 0·11 |
| **Highest education level** | |  |
| Mandatory school | 1 (Reference) |  |
| Vocational training/ baccalaureate | 0·74 (0·30 – 1·82) | 0·51 |
| Technical college or university | 0·70 (0·28 – 1·72) | 0·44 |
| **Employment status** | |  |
| Employed | 1 (Reference) |  |
| Student | 1·28 (0·74 – 2·20) | 0·87 |
| Unemployed | 0·89 (0·33 – 2·41) | 0·82 |
| **App notification** | |  |
| Not app notified | 1 (Reference) |  |
| App notified | 1·72 (1·25 – 2·36) | 0·0008 |
| *AHR: adjusted hazard ratio, CI: confidence interval*  *^*^Model adjusted for age group, sex, highest education level, employment status and app notification* | | |

***Model including only those reporting non-household exposure setting:***

| **Variable** | **AHR (95% CI) ^*^** | **p-value** |
| --- | --- | --- |
| **Age at diagnosis, years** | |  |
| 18-39 | 1 (Reference) |  |
| 40-64 | 1·26 (0·79 – 2·02) | 0·33 |
| 65+ | 1·24 (0·52 – 2·95) | 0·62 |
| **Sex** |  |  |
| Female | 1 (Reference) |  |
| Male | 0·79 (0·52 – 1·19) | 0·26 |
| **Highest education level** | |  |
| Mandatory school | 1 (Reference) |  |
| Vocational training/ baccalaureate | 1·07 (0·5 – 2·30) | 0·86 |
| Technical college or university | 1·04 (0·50 – 2·20) | 0·91 |
| **Employment status** | |  |
| Employed | 1 (Reference) |  |
| Student | 2·02 (1·00 – 4·06) | 0·05 |
| Unemployed | 2·20 (1·11 – 4·36) | 0·024 |
| **App notification** | |  |
| Not app notified | 1 (Reference) |  |
| App notified | 1·81 (1·22 – 2·68) | 0·003 |
| *AHR: adjusted hazard ratio, CI: confidence interval*  *^*^Model adjusted for age group, sex, highest education level, employment status and app notification* | | |

**Appendix 9: Sensitivity analysis using proxy exposure date instead of self-reported exposure date**

| **Variable** | **AHR (95% CI) ^*^** | **p-value** |
| --- | --- | --- |
| **Age at diagnosis, years** | |  |
| 18-39 | 1 (Reference) |  |
| 40-64 | 1·27 (0·90 – 1·79) | 0·17 |
| 65+ | 1·84 (0·86 – 3·93) | 0·11 |
| **Sex** |  |  |
| Female | 1 (Reference) |  |
| Male | 0·88 (0·64 – 1·19) | 0·40 |
| **Highest education level** | |  |
| Mandatory school | 1 (Reference) |  |
| Vocational training/ baccalaureate | 0·71 (0·36 – 1·42) | 0·33 |
| Technical college or university | 0·55 (0·25 – 1·19) | 0·13 |
| **Employment status** | |  |
| Employed | 1 (Reference) |  |
| Student | 1·38 (0·91 – 2·12) | 0·13 |
| Unemployed | 0·54 (0·23 – 1·26) | 0·15 |
| **App notification** | |  |
| Not app notified | 1 (Reference) |  |
| App notified | 1·36 (1·01 – 1·81) | 0·04 |
| *AHR: adjusted hazard ratio, CI: confidence interval*  *^*^Model adjusted for age group, sex, highest education level, employment status and app notification* | | |
